# What Do Persistent Misclassifications Tell Us About Alzheimer’s Disease Detection using Structural MRI?

**DOI:** 10.64898/2026.07.17.26358326

**Authors:** Didem Stark, Hwajin Shin, Nicolas Münster, Lydia Federmann, Kerstin Ritter, the Alzheimer’s Disease Neuroimaging Initiative

**Affiliations:** Hertie Institute for AI in Brain Health, University of Tübingen, Germany; Tübingen AI Center, Tübingen, Germany; The German Center for Mental Health Partner Site Tübingen (DZPG), Tübingen, Germany

**Keywords:** Alzheimer’s Disease, Disease subtyping, Deep learning, Misclassification analysis

## Abstract

Deep learning classifiers applied to structural MRI (sMRI) have achieved high performance in detecting Alzheimer’s Disease (AD), yet systematic investigation of their failure modes remains limited. In this study, we trained two deep learning architectures to classify AD from cognitively normal (CN) participants using sMRI data from the ADNI dataset, and examined whether misclassifications persist across models and training configurations. We identified a subgroup of subjects who were persistently misclassified across 100 model instances, and found that these subjects exhibited a markedly different atrophy subtype distribution compared to correctly classified AD cases, with substantial enrichment of hippocampal-sparing and minimal atrophy subtypes. To disentangle whether persistent false negatives (FN) reflect earlier disease stage or atypically presenting disease, we analyzed longitudinal follow-up scans and tested whether model predictions changed as neurodegeneration progressed. A change in prediction (from FN to true positive (TP)) was observed in only a subgroup of subjects and required intervals of up to five years, suggesting that persistent misclassification may not always be explained by disease staging alone. Although the sample size is small, these findings underscore the importance of accounting for disease heterogeneity in the development and evaluation of clinical AI models for AD detection.

## 1 Introduction

As the global burden of Alzheimer’s Disease (AD) accelerates, timely and accurate diagnosis becomes ever more critical [14]. Accordingly, recent years have seen a substantial increase in machine learning approaches to AD detection, particularly those leveraging structural magnetic resonance imaging (sMRI). Deep learning classifiers applied to sMRI can distinguish AD from cognitively normal (CN) controls with balanced accuracies exceeding 80% [12], yet a consistent finding across studies is that a subset of AD subjects is misclassified regardless of architecture or training configuration [9]. Kang et al. [9] showed, across seven architectures and multiple datasets, that false-negative (FN) cases cluster systematically: they are younger, less clinically impaired, and present with near-normal sMRI despite positive AD biomarkers, suggesting that model failures reflect the limits of structural neuroimaging as a disease marker rather than stochastic noise. This is mechanistically consistent with the amyloid/tau/neurodegeneration (A/T/N) framework [8, 7]: changes in grey matter volume reflect neurodegeneration, a late-stage event in the AD cascade that becomes detectable only after amyloid and tau pathology are already well established. Systematic classification errors of this kind may therefore encode clinically and neurobiologically meaningful heterogeneity rather than residual model failure [15].

AD is itself neuroanatomically heterogeneous at the individual level, with substantial variation in regional cortical-thickness abnormalities among patients, carrying the same clinical diagnosis [22]. Volume-based subtyping partitions AD subjects by age-, sex-, and intracranial-volume-adjusted hippocampal and cortical volumes into four canonical subtypes^4^: typical AD (tAD, ∼50.9%), limbic-predominant (LP, ∼19.4%), hippocampal-sparing (HpSp, ∼15.7%), and minimal atrophy (MA, ∼13.9%) [5]; we follow the classification algorithm of Risacher et al. [18], who defined the first three subtypes in ADNI, and explicitly integrate the MA subtype following Ferreira et al. [5]. The atypical subtypes, HpSp and MA, together account for roughly one third of amyloid-confirmed AD patients and share the absence of canonical medial temporal lobe atrophy: HpSp subjects exhibit rapid cognitive decline despite preserved hippocampal volumes [13], while MA subjects present with AD-level impairment despite relatively preserved brain structure [18].

Using 761 amyloid-confirmed ADNI subjects, we test whether persistent false-negative misclassification is associated with atrophy subtype. VoxCNN [4] and SFCN [16] were trained with repeated stratified cross-validation, and consensus errors were identified across architectures and initializations. We further applied the out-of-sample models to longitudinal follow-up scans. Persistent false negatives showed a distinct subtype distribution, underscoring the need for heterogeneity-aware evaluation of clinical AI models for AD detection.

## 2 Data, Methods, and Results

### 2.1 Data, Cohort, and Preprocessing

Data used in the preparation of this article were obtained from the Alzheimer’s Disease Neuroimaging Initiative (ADNI) database (adni.loni.usc.edu). The ADNI was launched in 2003 as a public-private partnership, led by Principal Investigator Michael W. Weiner, MD. The primary goal of ADNI has been to test whether serial magnetic resonance imaging (MRI), positron emission tomography (PET), other biological markers, and clinical and neuropsychological assessment can be combined to measure the progression of mild cognitive impairment (MCI) and early AD.

We included subjects from all ADNI phases who were cognitively normal (CN) or diagnosed with AD at baseline, with no diagnosis conversion at any later visit. Inclusion required a baseline 3T T1-weighted MRI. To reduce the risk of mislabeling, the AD group was restricted to amyloid-positive subjects and the CN group to amyloid-negative subjects. Amyloid status was determined from whichever assay was available, in the following priority order: (1) UPENN 2D-UPLC Mass Spectrometry (CSF), positive if A*β*42/40 *≤* 0.133; (2) UPENN Plasma Fujirebio/Quanterix, positive if ratio *≤* 0.0820; (3) C2N PrecivityAD2 Plasma, positive if APS2 *>* 47.5. One scan per subject was retained to prevent data leakage [24]. The final cohort comprised 761 subjects (575 CN; 186 AD).

Preprocessing comprised N4 bias field correction [21], skull-stripping with SynthStrip [6], non-linear registration to MNI152 space using ANTs [1], segmentation with SynthSeg [3], and WhiteStripe intensity normalisation [19].

### 2.2 Models, Training, and Analysis

VoxCNN is a 3D convolutional network with five convolutional blocks (channel progression 8 *→* 128), global average pooling, and a linear classifier [4, 11, 10]. Training used Adam (learning rate 1 *×* 10^*−*4^), cross-entropy loss, batch size 2, and early stopping (patience 8, maximum 200 epochs). SFCN [16], pre-trained on 14,503 UK Biobank scans for brain age estimation, was fine-tuned end-to-end for AD classification with all 2,950,466 parameters trainable (learning rate 5 *×* 10^*−*5^, softened class weights).

Training used a 5-fold × 4-repeat repeated stratified scheme (RepeatedStratifiedKFold), stratified by diagnosis, with a 70/10/20 train/validation/test ratio per fold. This yields 20 distinct splits, each trained with 5 random initializations, resulting in 100 trained models per architecture, with every subject appearing in the test set across all four repeats. Classification outcomes were aggregated across all splits, runs, and both architectures. A subject received a voted label (Voted-TP, Voted-TN, Voted-FP, Voted-FN) if at least 80% of evaluations agreed, and was labeled Mixed otherwise. Clinical, biomarker, and genetic variables (CDR-SB, ADAS-Cog 13, A*β*42/40, pTau217, and APOE *ε*4) were compared across voted groups using Mann–Whitney U tests.

AD subjects were assigned to Risacher subtypes [18] via a 2 *×* 2 median split of age-, sex-, field-strength-, and intracranial volume-adjusted hippocampal volume (HV) and cortical total volume (CTV), using ADNI cross-sectional FreeSurfer ROI data (UCSFFSX7). Adjusted cut-off equations were taken directly from Risacher et al. [18], who applied the same method to ADNI. Where the exact scan was unavailable, the closest scan from the same subject was used (median gap: 0 days; 86.6% within 30 days); subjects exceeding 30 days were excluded. An anatomical quality filter removed implausible HV:CTV ratios outside [0.05, 0.40], excluding approximately 11% of observations. Subtypes were: typical AD (tAD: low HV, low CTV), limbic-predominant (LP: low HV, high CTV), hippocampal-sparing (HpSp: high HV, low CTV), and minimal atrophy (MA: high HV, high CTV). Following Ferreira et al. [5], the MA subtype was explicitly integrated to distinguish the subjects in whom both HV and CTV were minimally affected. The resulting subtype distribution naturally varies from reported meta-analytic pools as they have pooled both AD-only and AD-and-CN quantification methods in structural AD subtyping. Subtype composition was compared between Voted-FN and Voted-TP using Fisher’s exact test with Cramér’s *V* .

To test whether false-negative misclassification reflects earlier disease stage rather than a persistent atypical presentation [17], later ADNI scans were identified for Voted-FN subjects. Only model instances that had originally evaluated each subject out-of-sample were applied, preserving the integrity of the out-of-sample evaluation. A minimum interval of 10 days between baseline and follow-up was required.

### 2.3 Results

#### Cohort

AD subjects were older (74.4 ±8.1 vs 69.1 ± 6.7 years) and more often male (52.2% vs 38.1%) than CN subjects. Risacher subtyping was applied to the 172 of 186 AD subjects with FreeSurfer ROI data, yielding 38 tAD (22.1%), 48 LP (27.9%), 48 HpSp (27.9%), and 38 MA (22.1%). The distribution departs from uniform because adjusted hippocampal and cortical volumes are negatively correlated in this sample, inflating the off-diagonal subtypes (LP, HpSp) (Table 1).

**Table 1.** Cohort and AD subtype demographics. Values are mean ± SD or percentage. Subtypes are the Risacher classification of the 172 of 186 AD subjects with FreeSurfer ROI data.

| Group | $n$ | Age | Female% | AD Subtype | $n$ | Age | Female% |
| --- | --- | --- | --- | --- | --- | --- | --- |
| AD | 186 | $74.4 \pm 8.1$ | 47.8 | tAD | 38 | $74.3 \pm 8.6$ | 34.2 |
| CN | 575 | $69.1 \pm 6.7$ | 61.9 | LP | 48 | $75.5 \pm 6.6$ | 75.0 |
| | | | | HpSp | 48 | $70.8 \pm 8.1$ | 33.3 |
| | | | | MA | 38 | $77.8 \pm 7.5$ | 39.5 |

#### Classification Performance

Both architectures discriminated AD from CN well (Table 2). SFCN outperformed VoxCNN on every metric: balanced accuracy 91.4% vs 88.0%, AUC 98.0% vs 95.6%. Sensitivity was below specificity for both models (VoxCNN 83.2% vs 92.8%; SFCN 88.2% vs 94.6%). At the 80% consensus threshold, the 761 subjects distributed as 524 Voted-TN, 146 Voted-TP, 13 Voted-FN, 11 Voted-FP, and 67 Mixed (27 AD and 40 CN without consensus).

**Table 2.** Classification performance (%± SD), averaged across 100 model instances per architecture.

| Model | Balanced Acc. | Sensitivity | Specificity | AUC |
| --- | --- | --- | --- | --- |
| VoxCNN | $88.0 \pm 3.2$ | $83.2 \pm 7.6$ | $92.8 \pm 4.0$ | $95.6 \pm 1.9$ |
| SFCN | $91.4 \pm 3.1$ | $88.2 \pm 7.3$ | $94.6 \pm 3.7$ | $98.0 \pm 1.0$ |

#### Persistent Misclassifications

Voted-FN subjects were younger than Voted-TP subjects (median 68 vs 76 years; *p* = 0.0039) and female-majority (61.5%), despite the AD cohort being approximately sex-balanced. Voted-FP subjects were the oldest group (77.5 ± 5.3 years) and predominantly male (72.7%). Cognitive severity followed the same separation: Voted-FN subjects were impaired but less so than Voted-TP, with lower CDR-SB (3.5 vs 4.5; *p* = 0.0082) and ADAS-Cog 13 (26.3 vs 31.7; *p* = 0.0047), while Voted-FP subjects were functionally unimpaired (CDR-SB median 0) but showed elevated ADAS-Cog 13 relative to Voted-TN. Plasma biomarkers and APOE *ε*4 status did not separate Voted-FN from Voted-TP: A*β*42/40 was comparable (0.077 vs 0.080; *p* = 0.68), as was p-tau217 (0.511 vs 0.614 (pg/mL); *p* = 0.50), and APOE *ε*4 positivity was high in both groups (66.7% vs 70.5%; *p* = 0.75) (Table 3).

**Table 3.**
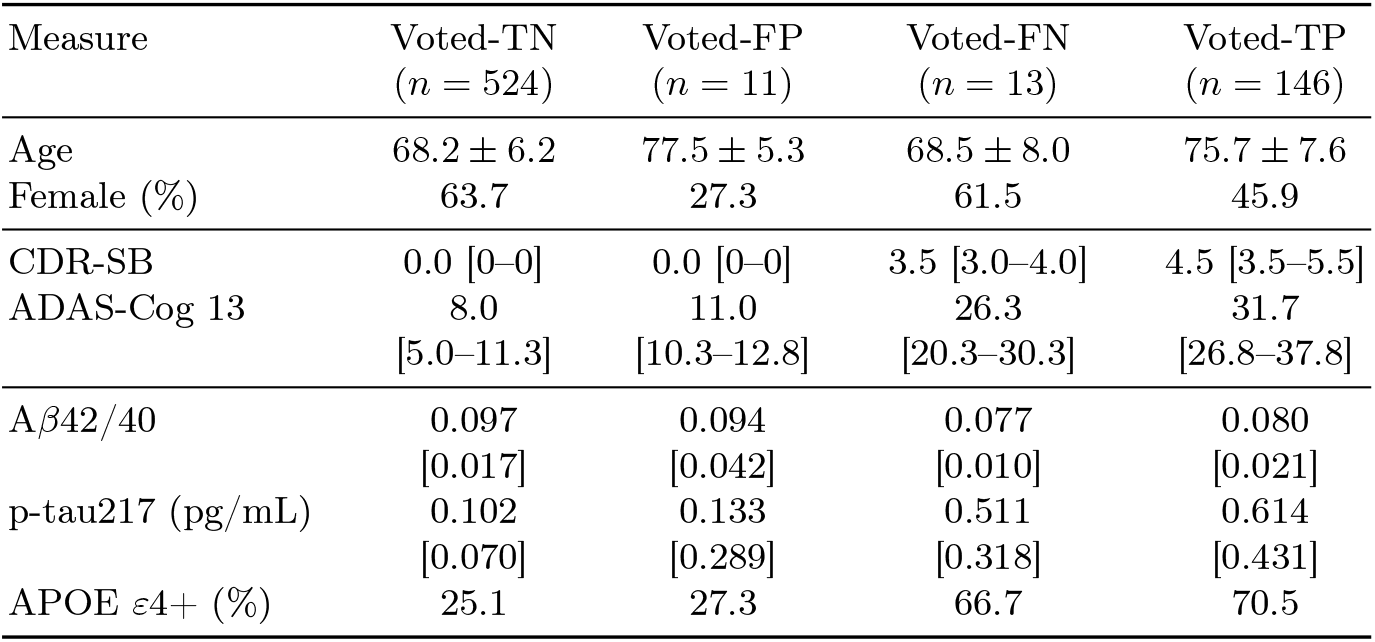
Demographic, clinical, biomarker, and genetic profiles by voted category. Age is mean± SD; sex is percentage; clinical scores are median [Q1–Q3]; plasma biomarkers are median [IQR]; APOE *ε*4 is percentage positive. The Mixed group (*n* = 67) is omitted.

| Measure | Voted-TN<br>($n = 524$) | Voted-FP<br>($n = 11$) | Voted-FN<br>($n = 13$) | Voted-TP<br>($n = 146$) |
| --- | --- | --- | --- | --- |
| Age | $68.2 \pm 6.2$ | $77.5 \pm 5.3$ | $68.5 \pm 8.0$ | $75.7 \pm 7.6$ |
| Female (%) | 63.7 | 27.3 | 61.5 | 45.9 |
| CDR-SB | 0.0 [0–0] | 0.0 [0–0] | 3.5 [3.0–4.0] | 4.5 [3.5–5.5] |
| ADAS-Cog 13 | 8.0<br>[5.0–11.3] | 11.0<br>[10.3–12.8] | 26.3<br>[20.3–30.3] | 31.7<br>[26.8–37.8] |
| Aβ42/40 | 0.097<br>[0.017] | 0.094<br>[0.042] | 0.077<br>[0.010] | 0.080<br>[0.021] |
| p-tau217 (pg/mL) | 0.102<br>[0.070] | 0.133<br>[0.289] | 0.511<br>[0.318] | 0.614<br>[0.431] |
| APOE ε4+ (%) | 25.1 | 27.3 | 66.7 | 70.5 |

The Risacher subtype composition differed significantly between Voted-FN (*n* = 12) and Voted-TP (*n* = 136) subgroups (Table 4). The Voted-FN group exhibited a higher proportion of HpSp (58.3% vs 21.3%) and MA (25.0% vs 22.1%) configurations alongside a distinct depletion of tAD (8.3% vs 26.5%) and LP (8.3% vs 30.1%) phenotypes. Overall, atypical subtypes comprised 83.3% of false negatives against 43.4% of true positives (*χ*^2^ = 9.552, *p* = 0.023, Cramér’s *V* = 0.254). See Figure 1 for the corresponding atrophy patterns.

**Table 4.**
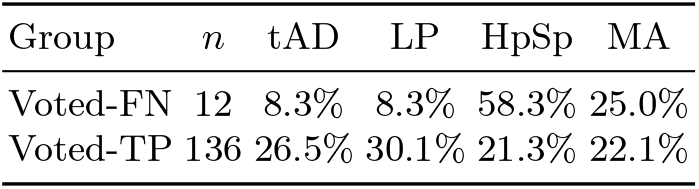
Risacher subtype composition by voted category (AD subjects with ROI data, unique subjects). Percentages are within-group.

**Fig. 1.**
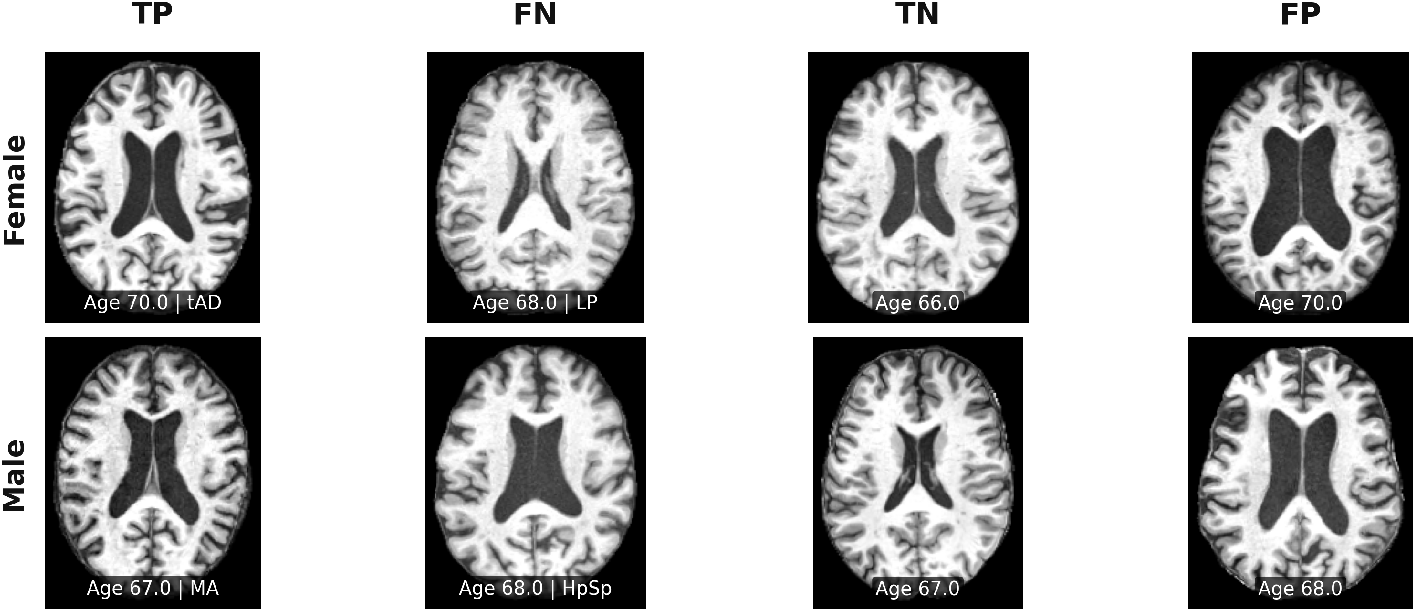
Representative T1-weighted MRI scans for one female and one male subject from each voted category. A single axial slice was selected at the midpoint of the volume along the z-axis. Columns show Voted-TP (true positive: AD correctly classified), Voted-FN (false negative: AD misclassified as CN), Voted-TN (true negative: CN correctly classified), and Voted-FP (false positive: CN misclassified as AD). Subject age and AD subtype where applicable are indicated below each panel.

Later scans were available for 9 of 12 Voted-FN subjects. Mean predicted AD probability rose from 21.6% at baseline to 36.8% at last follow-up, and 3 subjects crossed the decision threshold to Voted-TP (Table 5). Six of nine remained below threshold; subjects who experienced a longitudinal shift in predicted labels required evaluation intervals of up to 1,886 days; of three MA subjects, predicted label changed in only one.

**Table 5.**
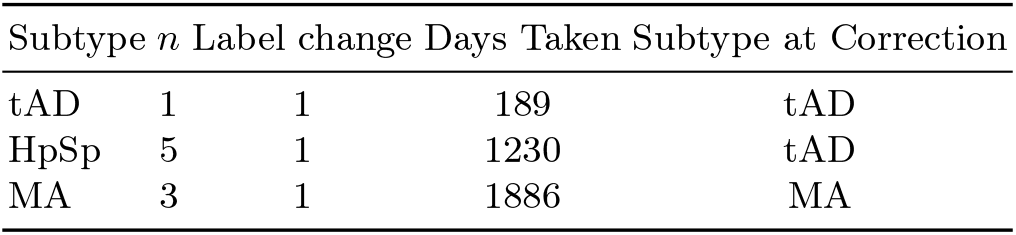
Longitudinal predicted label change of Voted-FN subjects by Risacher sub-type (subjects with later scans, *n* = 9). Correction denotes crossing the decision threshold to Voted-TP on any later scan.

## 3 Discussion

Across 100 model instances spanning two architectures, both models achieved high discriminative performance yet consistently failed on a distinct subset of AD subjects. Persistently misclassified false negatives were younger (median 68 vs. 76 years), more often female (61.5% vs. 45.9%), and showed markedly lower clinical severity on all cognitive measures despite comparable amyloid and tau burden, consistent with Kang et al. [9] and confirmed here under amyloid-verified inclusion criteria.

This study links persistent errors to atrophy subtypes. Voted false negatives were enriched for HpSp and MA subtypes (83.3% vs. 43.4%; *p* = 0.023, Cramér’s *V* = 0.254), while tAD and LP subtypes were substantially depleted. HpSp and MA share the absence of canonical medial temporal lobe atrophy. Thus, subjects carrying AD-level molecular pathology but preserving hippocampal and cortical volumes may be, from the model’s perspective, structurally indistinguishable from cognitively normal controls.

Whether persistent false negatives represent biologically atypical disease or simply earlier-stage disease not yet producing detectable atrophy is a non-trivial distinction. Our longitudinal follow-up of Voted-FN subjects (*n* = 9) offers some traction on this question. Mean predicted AD probability rose from 21.6% at baseline to 36.8% at last follow-up, and three subjects crossed the decision threshold; six of nine remained below it. These results support the idea that some persistent false negatives reflect early-stage disease that eventually converges toward a typical atrophy profile, while a substantial proportion remain structurally indistinguishable from cognitively normal controls even at later disease stages. False positives were the oldest group (mean 77.5 years), predominantly male (72.7%), and functionally unimpaired (CDR-SB median 0) despite mildly elevated cognitive scores, a profile consistent with age-related neurodegeneration morphologically resembling early AD but clinically buffered by cognitive reserve [20]

These findings should be interpreted in light of several limitations. ADNI recruits a predominantly white, highly educated, and medically monitored population, which likely underestimates structural heterogeneity in real-world clinical settings [2]. The longitudinal analysis is underpowered (*n* = 9 Voted-FN with follow-up), and subtype-stratified correction counts preclude statistical in-ference. The Risacher subtyping scheme is one of several available; data-driven longitudinal [17] or tau-PET-based approaches [23] may yield different subtype assignments. The 80% voted consensus threshold introduces a stability-versus-sample-size trade-off that warrants further exploration.

Persistent false negatives in structural MRI-based AD classification are systematically driven by atypical neuroanatomical phenotypes rather than stochastic noise. These failures are concentrated within hippocampal-sparing and minimal atrophy subtypes that harbor verified molecular pathology but lack canonical medial temporal lobe degeneration. Longitudinal data indicates that these errors reflect a combination of early-stage disease prior to structural convergence, alongside distinct trajectories that cross-sectional models cannot resolve from normal aging. Because these systematic misclassifications cluster within specific sub-populations defined by distinct age and clinical severity profiles, evaluating models purely on global performance metrics conceals critical failure modes. Accounting for individual structural heterogeneity is therefore a necessary prerequisite for building reliable diagnostic algorithms.

## Data Availability

All data used in this study is obtained from Alzheimer's Disease Neuroimaging Initiative (ADNI)
https://adni.loni.usc.edu/

https://adni.loni.usc.edu/

## Acknowledgments

This research was funded by the Deutsche Forschungsgemeinschaft (DFG) through the Walter Benjamin Programme (project number 565037378). Additional support was provided by the Gemeinnützige Hertie-Stiftung, the Tübingen AI Center, and the German Center for Mental Health (DZPG).

Data collection and sharing for this project was funded by the Alzheimer’s Disease Neuroimaging Initiative (ADNI) (National Institutes of Health Grant U01 AG024904) and DOD ADNI (Department of Defense award number W81XWH-12-2-0012). ADNI is funded by the National Institute on Aging, the National Institute of Biomedical Imaging and Bioengineering, and through generous contributions from the following: AbbVie, Alzheimer’s Association; Alzheimer’s Drug Discovery Foundation; Araclon Biotech; BioClinica, Inc.; Biogen; Bristol-Myers Squibb Company; CereSpir, Inc.; Cogstate; Eisai Inc.; Elan Pharmaceuticals, Inc.; Eli Lilly and Company; EuroImmun; F. Hoffmann-La Roche Ltd and its affiliated company Genentech, Inc.; Fujirebio; GE Healthcare; IXICO Ltd.; Janssen Alzheimer Immunotherapy Research & Development, LLC.; Johnson & Johnson Pharmaceutical Research & Development LLC.; Lumosity; Lundbeck; Merck & Co., Inc.; Meso Scale Diagnostics, LLC.; NeuroRx Research; Neurotrack Technologies; Novartis Pharmaceuticals Corporation; Pfizer Inc.; Piramal Imaging; Servier; Takeda Pharmaceutical Company; and Transition Therapeutics. The Canadian Institutes of Health Research is providing funds to support ADNI clinical sites in Canada. Private sector contributions are facilitated by the Foundation for the National Institutes of Health (www.fnih.org). The grantee organization is the Northern California Institute for Research and Education, and the study is coordinated by the Alzheimer’s Therapeutic Research Institute at the University of Southern California. ADNI data are disseminated by the Laboratory for Neuro Imaging at the University of Southern California.

## Disclosure of Interests

The authors have no competing interests to declare.

During the preparation of this manuscript, large language models (LLMs) were used for editing the text and debugging the code. All LLM-generated content was manually reviewed by the authors.

## Footnotes

4 Because the pooled subtype frequencies reported in [5] were estimated from overlapping but non-identical sets of studies and sum to 108%, we renormalized them to 100%.

